# Optimal seasonal timing of infant immunization to prevent RSV hospitalizations in Japan: a modelling study

**DOI:** 10.64898/2026.02.14.26346252

**Authors:** Ayaka Monoi, Akira Endo, Matej Kriznar, Motoi Suzuki, Stefan Flasche

## Abstract

The seasonal circulation of respiratory syncytial virus (RSV) in countries such as Japan, together with the transient nature of passive immunity conferred to infants via maternal vaccination or monoclonal antibody administration, may warrant a differential strategy for those born during the RSV inter-seasonal period. Maximal effectiveness may be achieved by deferring immunisation of this cohort from birth until entry into their first RSV season using catch-up administration of monoclonal antibody through a seasonal and catch-up programme, compared with year-round administration.

To estimate the benefit of seasonal and catch-up programmes in reducing RSV infant hospitalisations in Japan, we developed a static cohort model following infants through their first year of life, parameterised by Japanese data on weekly and municipality-specific RSV incidence during 2018 to 2025 and on RSV case hospitalisation risk from a health claims database study. We used Bayesian inference to estimate the effectiveness and its waning for maternal vaccine (RSVpreF) and long-acting monoclonal antibody (nirsevimab) from trial data.

We estimate that year-round programme of RSVpreF or nirsevimab could reduce RSV hospitalisations from the status quo, under which only high-risk infants are eligible for monoclonal antibodies, by 46% (95% uncertainty range (UR): 31%, 65%) or 58% (95%UR: 39%, 79%) respectively. Seasonal and catch-up programmes could achieve percentage reductions of 1.1-fold (95%UR: 0.82, 1.6) or 0.98-fold (95%UR: 0.83, 1.2) compared with the year-round programme. If seasonality matches the seasonal immunisation timing, using 2024 as an example, the percentage reduction was 1.2-fold (95%UR: 0.95, 1.6) or 1.1-fold (95%UR: 0.97, 1.2), respectively, compared with the year-round programme. If protection from nirsevimab remained substantial after six months, the year-round programme would likely to be more effective.

RSVpreF and nirsevimab may substantially reduce RSV infant hospitalisations in Japan. The benefit of the seasonal programmes depends on predictability of RSV seasonality and potential logistical challenges.

## Introduction

Respiratory syncytial virus (RSV) causes substantial pediatric morbidity and mortality worldwide. It is estimated that globally, RSV leads to approximately 100,000 deaths among children under 5 years old in a year. Particularly, young infants have a high risk of developing severe disease. RSV hospitalisation incidence among under-5-year-old in high-income countries is about 6 per 1,000 persons annually and that among infants aged 0-3-months is even higher at about 35 per 1,000 [1]. To protect infants against severe RSV disease, the World Health Organization’s (WHO) Strategic Advisory Group of Experts on Immunization (SAGE) recommended that all countries introduce the recently licensed immunisation products—maternal vaccine (RSVpreF) or long-acting monoclonal antibody (nirsevimab) [2]. Both products demonstrated sufficient efficacy in their clinical trials [3,4], although the protection from their passive immunisation is likely to wane within the first 6-12 months [4,5].

The magnitude of the benefit of administering RSV prophylaxes seasonally in combination with a catch-up administration of monoclonal antibody to infants born outside RSV season when they enter their first RSV season (“seasonal and catch-up programme”) is unclear. Instead of immunising irrespective of RSV season (“year-round programme”), in countries with strong seasonal RSV circulation, administering immunisation shortly before the RSV season may help ensure infants are optimally protected against the severe disease [6,7]. For instance, in the United States (U.S.) eligible individuals are recommended to receive either RSVpreF or monoclonal antibodies according to the local RSV season [8,9]. In contrast, some countries such as the United Kingdom (UK) administer RSVpreF as year-round administration. This approach may pose fewer logistical challenges than a seasonal and catch-up programme that combines seasonal immunisation for infants born during the RSV season and catch-up administration of monoclonal antibody for those born outside of the season [10,11].

Despite the potential benefits of seasonal RSV immunisation, identifying the optimal timing for administration remains a key challenge. While generally stable, RSV season can vary across regions within a country and may also change from year to year [2,12,13]. To maximise benefits at the subnational level, the Centers for Disease Control and Prevention (CDC) noted that public health authorities may elect to tailor administration timing in each area instead of applying a uniform schedule nationwide [8]. In addition to spatial heterogeneity, RSV circulation also exhibits temporal variability. RSV circulation was disrupted worldwide during the SARS-CoV-2 pandemic [14]. While some countries have reported that a return toward pre-pandemic RSV seasonality [15,16], Japan has not seen it as of 2025 [17]. The timing of RSV circulation in Japan has been changing both before and after the SARS-CoV-2 pandemic [17–19]. Currently proposed causes for the pre-pandemic shift include a change in proportion of the susceptible population [19], which might also occur in other settings. In addition, there is considerable heterogeneity in RSV seasonality across prefectures in Japan [17,20]. Under these conditions, implementing seasonal RSV immunisation that aligns well with the RSV season is challenging.

In 2025 November, the Japanese National Immunization Technical Advisory Group discussed that RSVpreF should be included in routine immunisation for pregnant women with gestation age of 28-36 weeks as year-round programme. Although the existing recommendation to limit the use of monoclonal antibodies to high-risk infants was upheld, the need for discussions on including nirsevimab in routine immunisation was recognised and it was noted that these discussions should be promoted swiftly, including whether the administration should be seasonal or year-round for infants aged under 1 year old [21]. Prior to the licensure of RSVpreF and nirsevimab, the only available prophylaxis was a short-acting monoclonal antibody, palivizumab. This is administered monthly to only high-risk infants, and in some prefectures the administration is during their first RSV season as seasonal and catch-up programme. Infants were eligible if, for instance, born before 29 weeks of gestational age and aged 12 months or younger at the beginning of the RSV season [22,23]. Due to the recent RSV seasonality that poses challenges to define an established RSV season, some prefectures have introduced year-round administration of palivizumab or nirsevimab for the high-risk infants depending on their eligibility [24–26].

This study evaluates potential impacts of RSV prophylaxes for all infants in Japan, focusing on the benefits of seasonal administration combined with catch-up administration of nirsevimab.

## Material and methods

We estimated the benefit of RSV infant immunisation programme using a static cohort model that follows infants through their first year of life. We then compared benefits between immunisation programmes using either RSVpreF or nirservimab and with different administration timings. To do so, we estimated RSV disease incidence by age in days and by calendar week using RSV surveillance data in Japan [17,27]. We estimated the waning efficacy of each product from efficacy data in their clinical trials [3,4]. We then estimated the percentage reduction in RSV under-1-year-old hospitalisations by an immunisation programme by comparing RSV hospitalisation incidence with and without that programme. Data analysis scripts used in this study are available at https://github.com/ayakamon/RSV_SEA_IMMUN.

This study used publicly available, anonymised data and therefore did not require ethical approval.

### Age-, prefecture- and time-dependent burden of RSV infant disease

To estimate time-dependent, age-dependent and prefecture-specific RSV hospitalisation rates, we multiplied estimated number of RSV hospitalisations for each calendar week in each prefecture by an estimated age distribution of all RSV hospitalisations in the dataset. The Japanese pediatric sentinel surveillance system reports laboratory-confirmed RSV cases on a calendar-week basis [27]. The number of sentinel sites was approximately 3,000 across the country until week 15 in 2025 and then reduced to about 2,000 thereafter due to a system reform [28]. To exclude the pandemic disruption to RSV transmission, we only considered the years 2018, 2019, 2023, 2024 and 2025 for this study. Age is grouped into 0-5 months, 6-11 months, and then by 1-year bands for 1-9 year old. Given that protection from the passive RSV immunisation may wane over the first few months, and severe RSV disease outcomes are likely among young infants, estimated impact of RSV infant immunisation may change depending on resolution of age groups of RSV disease. Thus, to obtain a disaggregated age distribution of RSV disease, we fitted a smoothing function following a gamma distribution to the age groups. Given that the age distribution of RSV cases is similar before the pandemic and 2023 and 2024 [17], we jointly fitted the single smoothing function to the age-grouped RSV cases in each prefecture in 2019 as the representative incidence. Given that the surveillance data does not include further information on severity beyond medically-attended cases, we then estimated the age distribution of RSV hospitalisation by multiplying the estimated age distribution of RSV cases by age-dependent RSV case hospitalisation risk derived from a study of health claim data in Japan [29]. We estimated age-dependent RSV hospitalisation incidence in each calendar week and prefecture by multiplying the estimated age distribution of RSV hospitalisation by temporally and spatially disaggregated incidence of RSV cases. Further details are described in SI1 and SI2.

### Defining the RSV season

In Japanese practice, the timing of future RSV immunisation is typically determined by past RSV incidence [30,31]. In this study the immunisation timing refers either to the calendar weeks when infants are born to vaccinated mothers or to the calendar weeks when infants receive nirsevimab. We defined the RSV seasonal immunisation timing in each prefecture as the continuous 26-week period during which the most RSV cases occurred in the past and hereafter we call such an immunisation strategy, which uses the prefecture-specific season, a prefecture-specific strategy. We considered scenarios where RSV season was defined based on a recent, typically the most recent, year reflecting the Japanese practice for monoclonal antibodies in some prefectures [30,31], or a more conservative scenario where the season was defined based on the weekly numbers of cases aggregated across multiple years. To define RSV season in this study, given the observed biannual RSV incidence in 2025, we used 2024 as the single reference year to define the RSV season instead of 2025. In the conservative scenario, we used 2018, 2019, 2023, and 2024 as reference years. We also estimated the impact of a seasonal strategy that defines the RSV season based on nationally aggregated data as a logistically more streamlined alternative to the prefecture-specific strategy for seasonal immunisation (hereafter referred to as nationally synchronised strategy).

### Vaccine effectiveness and waning protection following immunisation

We modelled the effectiveness of RSVpreF and nirsevimab by estimating their efficacy and waning from respective clinical trials [3,4] using a Bayesian framework. We assumed that the efficacy against RSV cases and hospitalisations would decline similarly with time since birth for RSVpreF or since immunisation for nirsevimab following an Erlang-2 distribution. We used Markov chain Monte Carlo methods to obtain the posterior distributions for the parameters that describe the waning protection for both products independently. Further details are described in SI3.

### Modelling the impact of vaccination

We estimated the impact of an immunisation programme offered to all infants regardless of their risk of developing severe RSV disease. We measured the impact as a reduction in RSV hospitalisations among infants aged less than 1 year from the status quo without RSVpreF or nirsevimab, but with palivizumab being used for high-risk infants. We calculated the reduction in RSV hospitalisations by prefecture as a product of the estimated age- and time-specific incidence of hospitalisation and effectiveness of RSVpreF or nirsevimab in preventing RSV hospitalisation over time, assuming 100% immunisation coverage. To obtain national estimates of reduction in RSV hospitalisations, we aggregated estimates across prefectures by weighing prefecture-specific estimates with the estimated number of RSV hospitalisations in each prefecture.

### Uncertainty

Given the uncertainty in seasonality of future RSV seasons, we assumed that the future season for which we estimate the impact of RSV prevention would be the same as one of the observations in 2018, 2019, 2023, 2024, or 2025. We then randomly sampled one annual incidence from the study years with replacement to account for uncertainty about future RSV seasonality in our estimates of impact of RSV immunisation programme. In addition, we considered scenarios where the seasonality matched that in 2024 or 2025. For each of the 10,000 iterations of calculation of reduction in RSV hospitalisation, we sampled (i) annual incidence of calendar-week dependent RSV hospitalisation (ii) immunisation effectiveness and (iii) age distribution of RSV disease. For each sampled annual incidence, we also sampled an effectiveness profile (effectiveness and its change in the first year either, since birth for RSVpreF or immunisation for nirsevimab) from their respective posterior distribution. We report the median and 95% uncertainty range (UR) as the 95% quantiles of the outcomes.

## Results

### Seasonality of RSV

In the four study years of 2018, 2019, 2023, and 2024, 120,000 – 145,000 RSV cases were reported annually across Japan. In the four study years, 76% of RSV cases were reported during the prefecture-specific RSV season. In comparison, 84% of RSV cases were reported within the 2024 RSV season. If defining a single national level RSV season over week 14-39, 74% of RSV cases were reported during RSV season across the four study years, and 82% in 2024. Within the country, the concentration of RSV activity into a prefecture-specific 6-month season varied from 56% of all cases reported in the season during the four years in Hokkaido, to 97% reported in the season in 2024 in Nagasaki. On the contrary, the percentage of RSV cases reported during a single national level season starting in week 14 varied from 55% of all cases over the four years in Hokkaido to 97% in 2024 in Nagasaki. The prefecture-specific seasonal immunisation timing in the four years varied from start week 10 in Okinawa (92% cases included) to week 26 in Nagano (69%). In contrast to the annual seasons in the other years, in 2025, two RSV peaks were observed at national level, as well as in most prefectures (Fig 1A and B). Thus, we did not include 2025 in defining the timing of seasonal administration.

**Figure 1.**
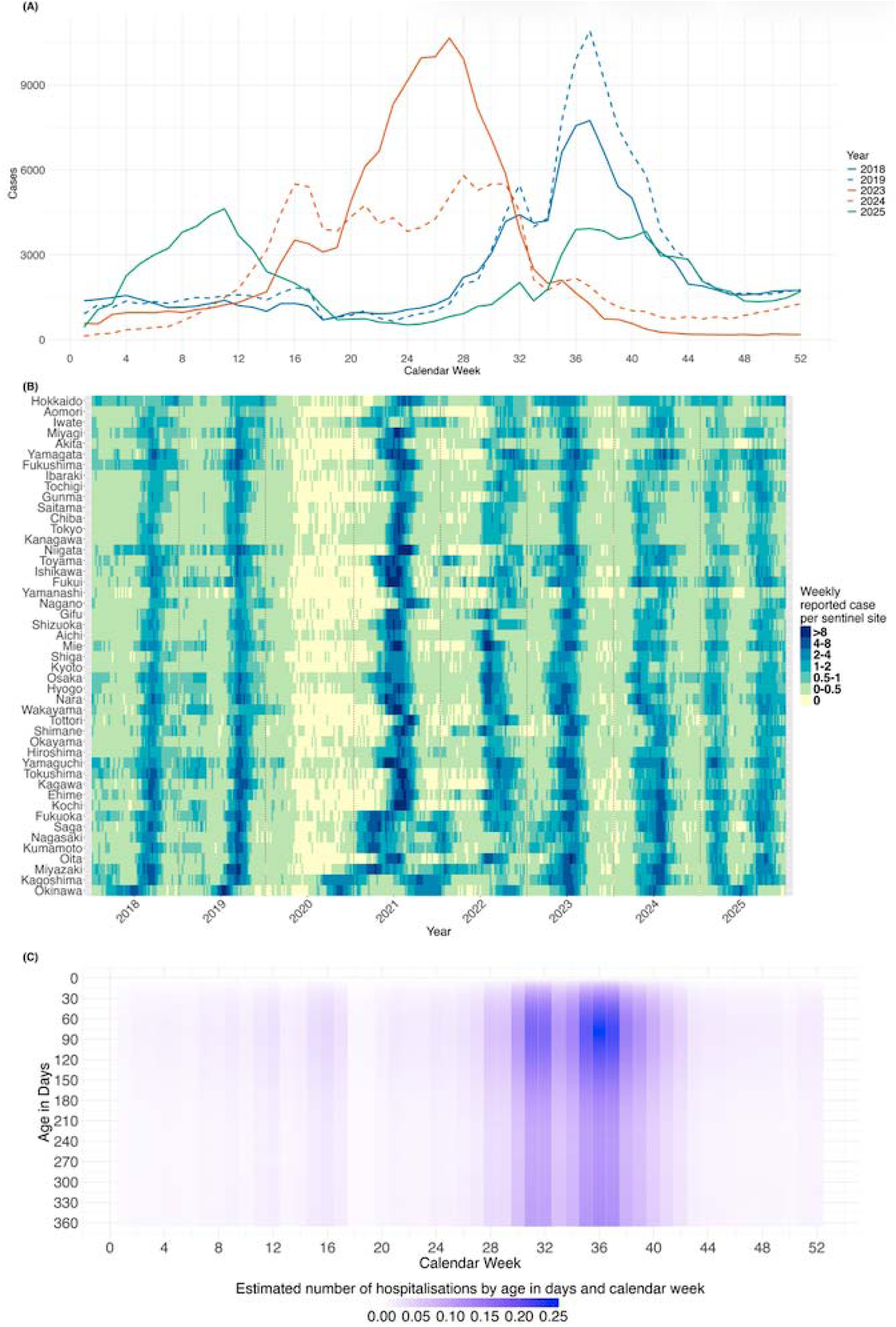
RSV incidence reported through the sentinel surveillance system in Japan. (A) RSV weekly reported cases at the national level in 2018, 2019, 2023, 2024 or 2025. (B) RSV weekly reported cases per sentinel site by prefecture in 2018-2025. Note that the sentinel surveillance system changed from week 15 in 2025. (C) Estimated number of RSV hospitalisations in a prefecture (here Tokyo, which has the largest population) in 2019 by age in days and by calendar week. The number of hospitalisations is shown using a purple gradient.

### The burden of RSV disease in Japan

We estimated a function representing the observed RSV notifications of any severity in Japan, with a peak incidence at 9 months of age, and that 50% of all under-10-year-old cases are from infants aged 3-19 months.

The RSV case hospitalisation risk in Japan declines rapidly with age. We estimated that 87% of cases among newborns are admitted to hospital whereas 22% of 6-month-old infants are admitted. However, the risk was suggested to be similar among infants aged from 6 months to 1 year (17-20%) (Fig S4).

We then estimated a function representing RSV hospitalisation incidence in Japan, with a peak incidence at 2 months of age with 50% of all hospitalisations among infants less than 1 year old occurring in 3-23 weeks of age (Fig 1C).

### Estimated effectiveness of immunisation

Based on the clinical trial results, modelled effectiveness of RSVpreF against severe RSV-associated MA-LRTIs was 87% (95% Crl: 67%, 99%) on the first day of life (Fig S6). Effectiveness was estimated to wane to 48% (95% Crl: 26%, 68%) at 150 days of age, and 10% (95% Crl: 1.5%, 45%) a year following birth, respectively.

Modelled effectiveness of nirsevimab against hospitalisation was 80% (95% Crl: 61%, 93%) on the first day after immunisation. Effectiveness was estimated to wane to 61% (95% Crl: 40%, 81%) at 150 days of age, and 29% (95% Crl: 8.0%, 70%) a year following birth, respectively.

### Estimated impact of RSV programmes

We estimated that, if all infants were immunised, year-round administration of RSVpreF or nirsevimab could reduce RSV infant hospitalisations from the status quo by 46% (95% UR: 31%, 65%) or 58% (95% UR: 39%, 79%), respectively (Table 1, Fig 2). For our baseline scenario of prefecture-specific seasonal administration of RSVpreF or nirsevimab combined with catch-up administration of nirsevimab, we estimated that 52% (95% UR: 37%, 65%) or 57% (95% UR: 38%, 75%) of hospitalisations could be prevented. This corresponded to 1.1 (95% UR: 0.82, 1.6) times of the percentage reduction in hospitalisations for the RSVpreF strategy, and to 0.98 (95% UR: 0.83, 1.2) times for the nirsevimab strategy.

**Table 1.**
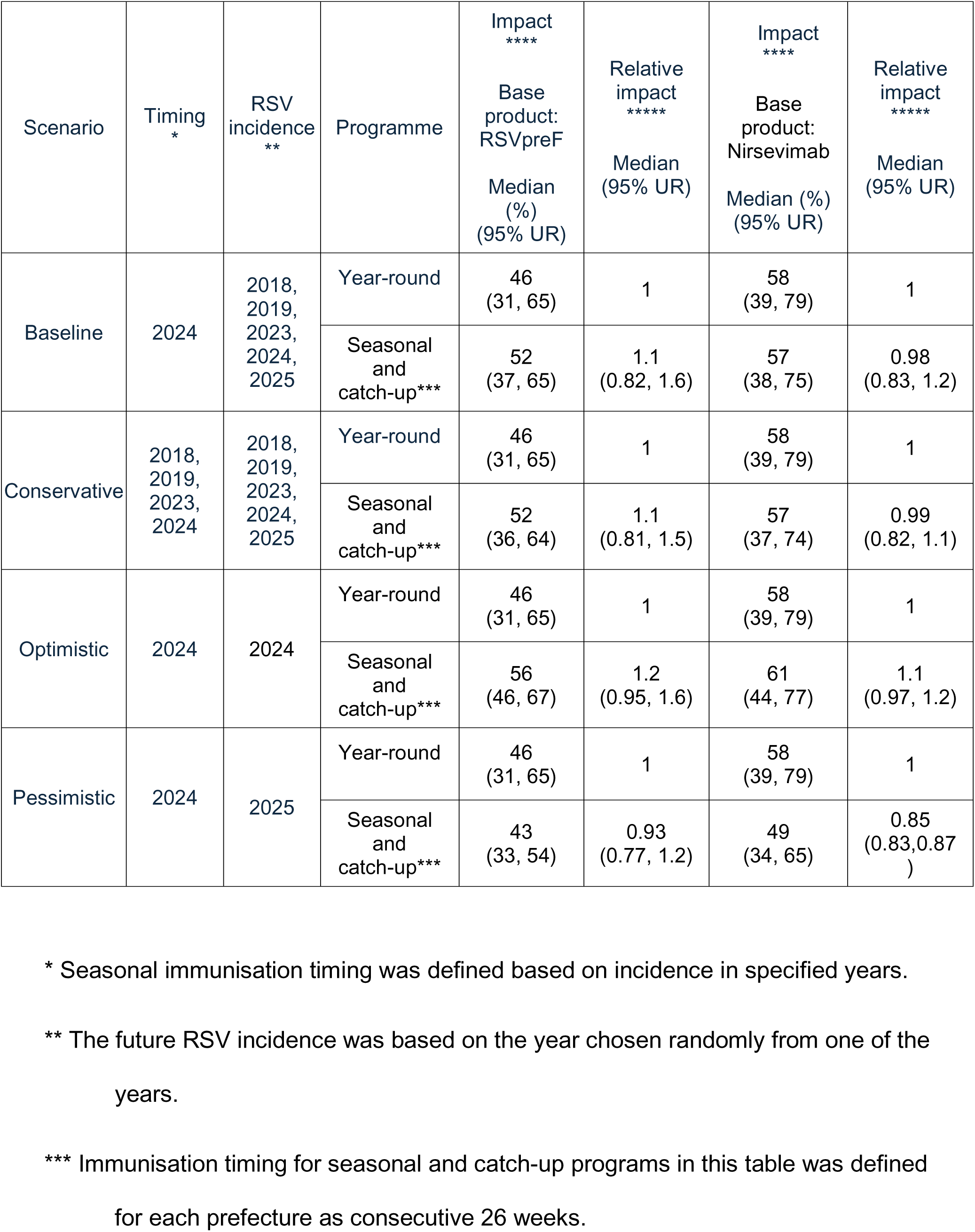
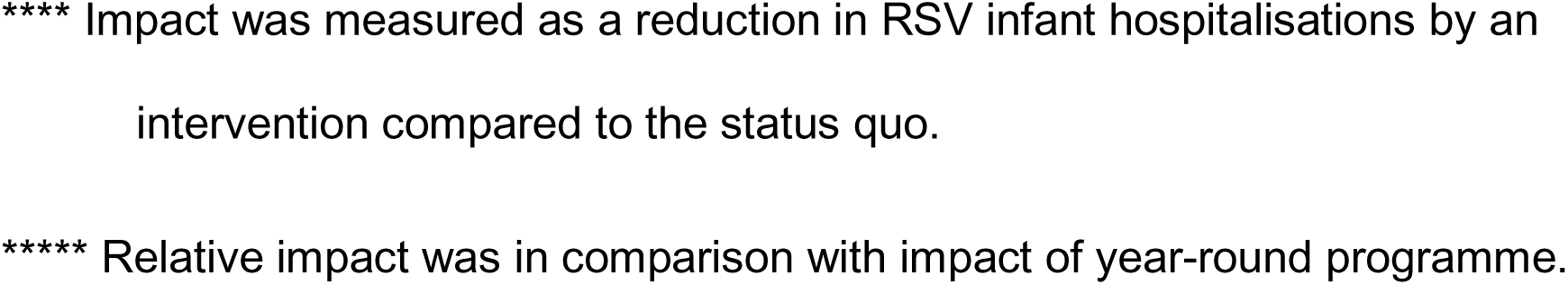
Estimated benefit of an RSV infant immunisation programme and additional benefit of seasonal administration compared to year-round administration by scenario. The benefit is a percentage reduction in under-1-year-old RSV hospitalisations by a programme compared to under-1-year-old RSV hospitalisations without that programme. Additional benefit is measured as relative increase in benefit between programs (i.e., seasonal and catch-up programme compared to year-round programme). We considered four scenarios depending on definition of immunisation timing, and RSV incidence for which we apply that immunisation. For each scenario, we estimated reduction in RSV infant hospitalisation for (i) each programme; i.e., year-round administration in all prefectures (yea-round programme), or seasonal administration with prefecture-specific timing combined with catch-up administration of nirsevimab for infants born outside RSV season (seasonal and catch-up programme), and also for (ii) product used for infants born during season (RSVpreF or nirsevimab). UR: uncertainty range.

**Figure 2.**
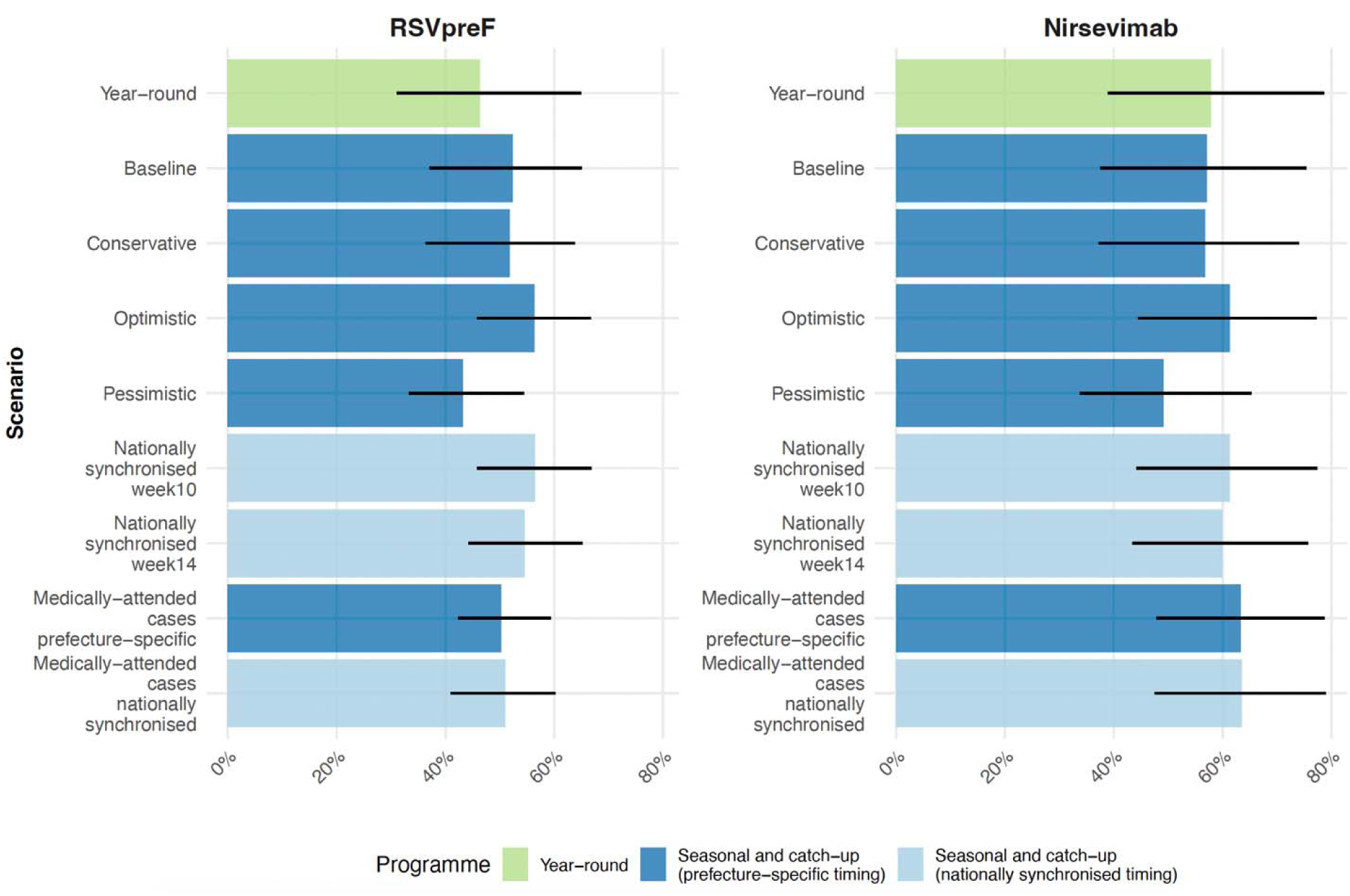
Estimated reduction in RSV infant disease outcomes by immunisation product and by scenario. The estimated reduction is a reduction in the number of under-1-year-old RSV hospitalisations from the status quo, unless it is stated as medically-attended cases. The immunisation product shown in each panel is a product given to the birth cohort. The bar charts colored in green show reductions by year-round programmes, and the bar charts colored in light or thick blue show reductions by seasonal and catch-up programmes. Nationally synchronised strategies start in week 10 or 14 if the outcomes are hospitalisations. If the disease outcomes are medically-attended cases, nationally synchronised strategies start in week 6 for RSVpreF or week 10 for nirsevimab, respectively. Further details of the scenarios of “Baseline”, “Conservative”, “Optimistic”, and “Pessimistic” are described in Table 1.

Similarly, as our conservative scenario, if seasonal immunisation timing was defined using the number of RSV cases aggregated over the four study years, we estimated that the prefecture-specific seasonal administration would reduce 1.1-fold (95% UR: 0.81, 1.5) RSV hospitalisations for the RSVpreF strategy, and 0.99-fold (95% UR: 0.82, 1.1) for the nirsevimab strategy. As our optimistic scenario, if seasonal immunisation timing aligned perfectly with actual season (using 2024 epidemiology as an example), the impact of the seasonal and catch-up programme would increase to 1.2 (95% UR: 0.95, 1.6) times compared to that of the year-round strategy for RSVpreF and by 1.1 (95% UR: 0.97, 1.2) times for nirsevimab. However, as our pessimistic scenario, if seasonal immunisation timing was poorly matched to the season (i.e., if we use immunisation timing based on the 2024 season and future season is similar to 2025), seasonal immunisation may prevent substantially fewer hospitalisations than year-round immunisation. We estimated that, in this case, the seasonal programme may prevent 0.93 (95% UR: 0.77, 1.2) and 0.85 (95% UR: 0.83, 0.87) times as many RSV hospitalisations.

In analysing the estimates, we found that the added benefit of seasonally focused immunisation was strongly correlated with the estimated durability of protection following immunisation, and that the correlation was stronger if the RSV season matched well with seasonally delivered immunisation (Fig S10). In the baseline scenario, the benefit of a seasonally focused programme would outweigh that of year-round programme, if the effectiveness of nirsevimab declined in the first 180 days after administration by more than 10% in 2023 to 52% in 2018. However, if RSV epidemiology was the same as 2025, the year-round programme would outweigh the seasonal and catch-up programme regardless of the durability of the effectiveness.

### Scenario analyses

We considered scenario analyses varying the potential key factors on the impact, namely, immunisation coverage, severity of target RSV disease outcome, prefectures, and having nationally synchronised timing of seasonal administration across prefectures or prefecture-specific timing.

#### Potentially reduced immunisation coverage

Given a potential lower coverage if delivered as a catch-up cohort for part of the birth cohort, we assessed how robust the gains from seasonal immunisation would be to the coverage. For the RSVpreF-based programme in the optimistic scenario, we estimated that if the immunisation coverage among those eligible for catch-up immunisation in the seasonal programme is at least 69% of that achieved by administration of RSVpreF, the seasonal programme would have a larger impact.

For the nirsevimab-based programme, we estimated that coverage among the catch-up eligible infants will need to exceed 90% of that achieved through immunising at birth in order to increase impact through seasonal administration.

#### Nationally synchronised seasonal administration

We explored how effective a seasonal strategy that defines the RSV season based on nationally aggregated data would be as a logistically more streamlined alternative to the prefecture-specific strategy for seasonal immunisation. Assuming the future RSV incidence would be the same as observed RSV incidence in 2024, we estimated that such national synchronised strategy of seasonal administration of RSVpreF or nirsevimab combined with catch-up administration of nirsevimab (seasonal and catch-up programme) starting in week 10 would best reduce RSV hospitalisations by 56% (95% UR: 46%, 67%) or by 61% (95% UR: 44%, 77%), respectively. In comparison, if the six-month immunisation starts 4 weeks later, the seasonal and catch-up programme (starting in week 14) could reduce 55% (95% UR: 44%, 65%) or 60% (95% UR: 43%, 76%), respectively (Fig 3, Fig S11).

#### Regional heterogeneity in impact

We estimated that the impact of year-round immunisation with RSVpreF or nirsevimab was the same across prefectures in reducing RSV hospitalisations (46% with RSVpreF or 58% with nirsevimab). In our baseline scenario, our estimated impact under seasonal administration showed spatial heterogeneity favoring prefectures with highly concentrated RSV epidemics: we find that the impact of strategy with RSVpreF or nirsevimab ranged from a 45% (95% UR: 32%, 62%) and 51% (95% UR: 33%, 72%) reduction in RSV hospitalisations in Hokkaido to 57% (95% UR: 46%, 68%) and 62% (95% UR: 45%, 78%) in Okinawa. We find similar heterogeneity in impact under nationally synchronised seasonal administration. Further details are described in SI7.

#### Disease severity

We also estimated that defining prefecture-specific seasonal immunisation timing would bring less additional benefits in reducing less severe RSV disease outcomes. In our optimistic scenario, RSV-preF or nirsevimab-based prefecture-specific strategy could reduce medically-attended RSV cases by 50% (95% UR: 42%, 59%) or 63% (95% UR: 48%, 79%), respectively. On the contrary, we estimated that, if assuming the future RSV incidence is the same as the observations in 2024, a single national strategy in which infants born in week 6-31 are born to mothers who have received RSVpreF would best reduce infant medically-attended RSV cases by 51% (95% UR: 41%, 60%). For nirsevimab national strategy with immunisation during week 10-35 would best reduce by 64% (95% UR: 47%, 79%).

## Discussion

By using the country-specific RSV surveillance and hospitalisation risk estimates, we find that both RSVpreF and nirsevimab have the potential to reduce RSV infant hospitalisation in Japan substantially. We show that seasonal administration combined with catch-up administration for those born outside the RSV season has the potential to reduce the burden of RSV beyond what is possible through year-round immunisation. However, the potential gain from seasonal administration strongly depends on the ability to predict RSV seasonality for each prefecture and the ability to achieve high coverage among those eligible to receive immunisation as part of a catch-up.

Our impact predictions align with emerging evidence on effectiveness of RSV immunisation from countries where administration has started. For instance, the CDC recommends seasonal administration of either of RSVpreF or long-acting monoclonal antibodies for eligible individuals; for infants born outside RSV season, catch-up administration of long-acting monoclonal antibody is recommended at the beginning of the seasonal administration window [8]. As of February 2025, an estimated 66% of the infants received one of the products and an estimate using data from U.S. surveillance systems suggests a 43-56% reduction of RSV hospitalisations among infants aged 0-7 months in the 2024-2025 RSV season in locations where RSV prophylaxes were widely available before the RSV season [32]. In Galicia, Spain, about 95% of eligible infants received nirsevimab during the 2023-24 campaign and in the 2023-24 RSV season the estimated reduction in RSV-related LRTI hospitalisations was 85.9% (95% CI: 80.2%, 90.0%) and 55.3% (95% CI: 22.5%, 74.3%) in the 2024-25 season [33].

Whether RSV immunisation is offered seasonally varies substantially across countries; for instance, year-round administration is implemented in the UK and Australia, whereas seasonal administration is implemented in countries such as Argentina, Germany, France, Austria, Belgium, and U.S. [7,11]. While country-specific RSV seasonality and local logistical context need to be considered, further evidence is needed to inform countries that are currently deliberating the introduction of new RSV immunisation programmes.

The past 10 years of RSV circulation in Japan put into question our ability to accurately predict the timing of the upcoming season in order to issue seasonal immunisation guidance. In addition to the marked differences between pre- and post-pandemic RSV seasons [17], the distinct biannual peaks in RSV notification observed in 2025 complicates the implementation of seasonal administration strategies. Biannual RSV peaks have been previously reported using surveillance data in Hong Kong prior to the COVID-19 pandemic [34]. Since the onset of the pandemic, biannual peaks of RSV infections have also been reported in 2022 from a hospital-based study in China [35].

However, the factors underlying the distinct biannual RSV peaks in Japan in 2025 remain unknown, although they may include changes in contact behavior, demographic changes or immunity dept from the lack of exposure during pandemic years. If logistically feasible, one could devise a responsive immunisation strategy that is triggered by an incidence threshold [36]. However, even under such a strategy, the relatively short interval between the two RSV seasons in 2025 would likely challenge the effective delivery and may not differ much from year-round administration.

The achievable coverage of the new RSV infant programmes in Japan is uncertain. RSVpreF is the first maternal vaccine recommended for inclusion in the national immunisation programme in Japan and a monoclonal antibody programme for healthy infants that includes catch-up administration would also represent a novelty. While the coverage of routine immunisation in infants in Japan is high [37], the acceptance of both maternal immunisation and immunisation shortly after birth will need to be explored. A survey between July 2024 and August 2025 suggests that among 1279 respondents 11.6% had received maternal RSV vaccination and 77.5% of the respondents answered that they would accept vaccination only if no out-of-pocket payment were required [38]. Coverage in countries where RSVpreF has already been introduced has been in a similar line with maternal immunisation coverage for pertussis [39,40] but remained substantially below the levels typically observed for routine childhood immunisation. For instance, as of May 2025, 54.0% of eligible mothers in the UK received RSVpreF under a year-round programme [41], while in Argentina national coverage reached 62.5% among pregnant women during the first season by August 2024 under a seasonal programme [40]. In comparison, coverage for nirsevimab may be higher in some countries; for instance, cumulative coverage up to 30 March 2025 in Galicia, Spain, was reported on 2 April 2025 to be 90.7% at birth and 89.4% in the catch-up group [42]. In France, for the 2024-2025 season the guideline recommended either maternal vaccination or nirsevimab during the first year of life. In the second year since the national immunisation programme with nirsevimab started, the coverage of nirsevimab has been estimated at 84% at maternity wards and 60% if born outside the RSV season. Our findings suggest that for seasonally focused programmes to outperform year-round programmes, high coverage in the catch-up group will be required. Context-specific and feasible strategies may therefore be needed to increase catch-up coverage [43].

We show that the risk of RSV hospitalisation is highest earlier in life than the peak of medically-attended RSV cases. As a result, the estimated benefit of seasonally focused administration is higher for our more severe clinical outcome, because it prioritises protection in early infancy. Our estimated peak in age distribution of RSV hospitalisation is similar to that in a health claim database study in Japan [29]. In this study we focus exclusively on the impact on RSV hospitalisations due to the lack of data on more severe outcomes; however, the burden of more severe outcomes is concentrated even earlier in life and thus seasonal administration may offer some additional benefit beyond what we estimated for averting RSV hospitalisations.

We show that the additional benefit of a seasonal and catch-up programme compared with a year-round programme is strongly correlated with the waning of protection following immunisation; faster waning favours seasonal and catch-up programmes.

Currently only a limited number of studies provide evidence on the duration of protection. Our trial-based estimates for the effectiveness of nirsevimab are consistent with findings from a test-negative case-control study in three European countries [44]. However, a test-negative case-control study in the United States reported more rapid waning, with the effectiveness of nirsevimab against RSV hospitalisation declining from 90.6% (95% Crl: 70.7, 98.3) to 48.8% (95% Crl: -149, 87.5) over 16 weeks following immunisation [5]. Conversely, an effectiveness study in Chile suggests that protection against RSV-related lower respiratory tract infection hospitalisations may last beyond 90 days [45].

Clesrovimab, another long-acting monoclonal antibody recently licensed in countries such as the U.S., may provide a different duration and hence the benefit of seasonal programme may change [46]. However, evidence on protection beyond six months remains scarce, as does that for RSVpreF.

There are some key limitations to our study. Firstly, given that the peak age of RSV infant hospitalisation is older than the time of their birth, there may be a time lag between our definition of prefecture-specific seasonal immunisation timing - defined as a six-month period intended to capture the majority of RSV cases - and the optimal timing for reducing RSV hospitalisations among infants. As a result, we may underestimate the benefits of seasonal administration. However, by varying seasonal immunisation timing, we also show that the difference between the best impacts and the impacts achieved by the defined seasonal immunisation may not be substantial.

Moreover, alternative definitions of RSV season may be applicable. Different methods to define the RSV season could also be considered [6] [47], however, our estimates are in line with the current practice of countries where RSV immunisation is implemented [40,48,49]. We also did not assess impacts on more severe outcomes such as deaths nor did we assess the higher risk of hospitalisation among children with commodities. While a key objective of RSV infant immunisation is to prevent severe RSV disease particularly among young infants who are at high risk, the societal impact of less severe but symptomatic RSV infections in children may also be substantial [50]. Finally, we did not consider the potential difference in the prices of the products in this analysis.

## Conclusion

Both RSVpreF and nirsevimab can substantially reduce the burden of RSV associated hospitalisations in infancy in Japan. The impact of the programme may be improved by about 1.1-1.2 times through seasonal administration but this hypothetical gain may erode by changing seasonality, reduced coverage in those eligible for catch-up immunisation, or better-than-anticipated durability of protection.

## Declaration of generative AI and AI-assisted technologies in the manuscript preparation process

During the preparation of this work the authors used ChatGPT to improve readability and correct grammatical inaccuracies in the draft. After use, the authors reviewed and edited the content as needed and take full responsibility for the content of the published article.

## Contributions

Conceptualization; AM, SF, MS

Data curation; AM

Formal analysis; AM

Funding acquisition; AM

Investigation; AM

Methodology; AM, SF, MS, AE

Project administration; AM, MS

Resources; AM, MS

Software; AM, MK

Supervision; SF, MS, AE

Validation; AM, SF, MS, AE, MK

Visualization; AM

Roles/Writing - original draft; AM

Writing - review & editing; AM, SF, MS, AE, MK

All authors provided final approval of the version to be published and agreement to be accountable for all aspects of the work in ensuring that questions related to the accuracy or integrity of any part of the work are appropriately investigated and resolved.

## Data Availability

All data produced are available at

## Acknowledgements

None.

## Competing interests

The authors declare no conflict of interest.

## Funding

AM was supported by the Japanese Ministry of Education, Culture, Sports, Science and Technology through the Doctoral Program for World-leading Innovative & Smart Education as part of the NU-LSHTM Joint PhD Programme for Global Health. AE was supported by Japan Science and Technology Agency (JPMJPR22R3), the Japan Society for the Promotion of Science (JP22K17329) and Japan Agency for Medical Research and Development (JP223fa627004). SF was supported by the Einstein Foundation (EPP-BUA-2022-697). The funders had no role in study design, data collection and analysis, decision to publish, or preparation of the manuscript.

## Notes

### Competing Interest Statement

The authors have declared no competing interest.

### Author Declarations

The study used (or will use) ONLY openly available anonymized data that were originally located at:the website of National Institute of Infectious Diseases, Japan.

