## Supplemental information for "Optimal seasonal timing of infant immunization to prevent RSV hospitalizations in Japan: a modelling study"

### Supporting information

#### 1 Overview of the model

We developed a static cohort model that is disaggregated spatially and temporally specifically for RSV disease burden in Japan.

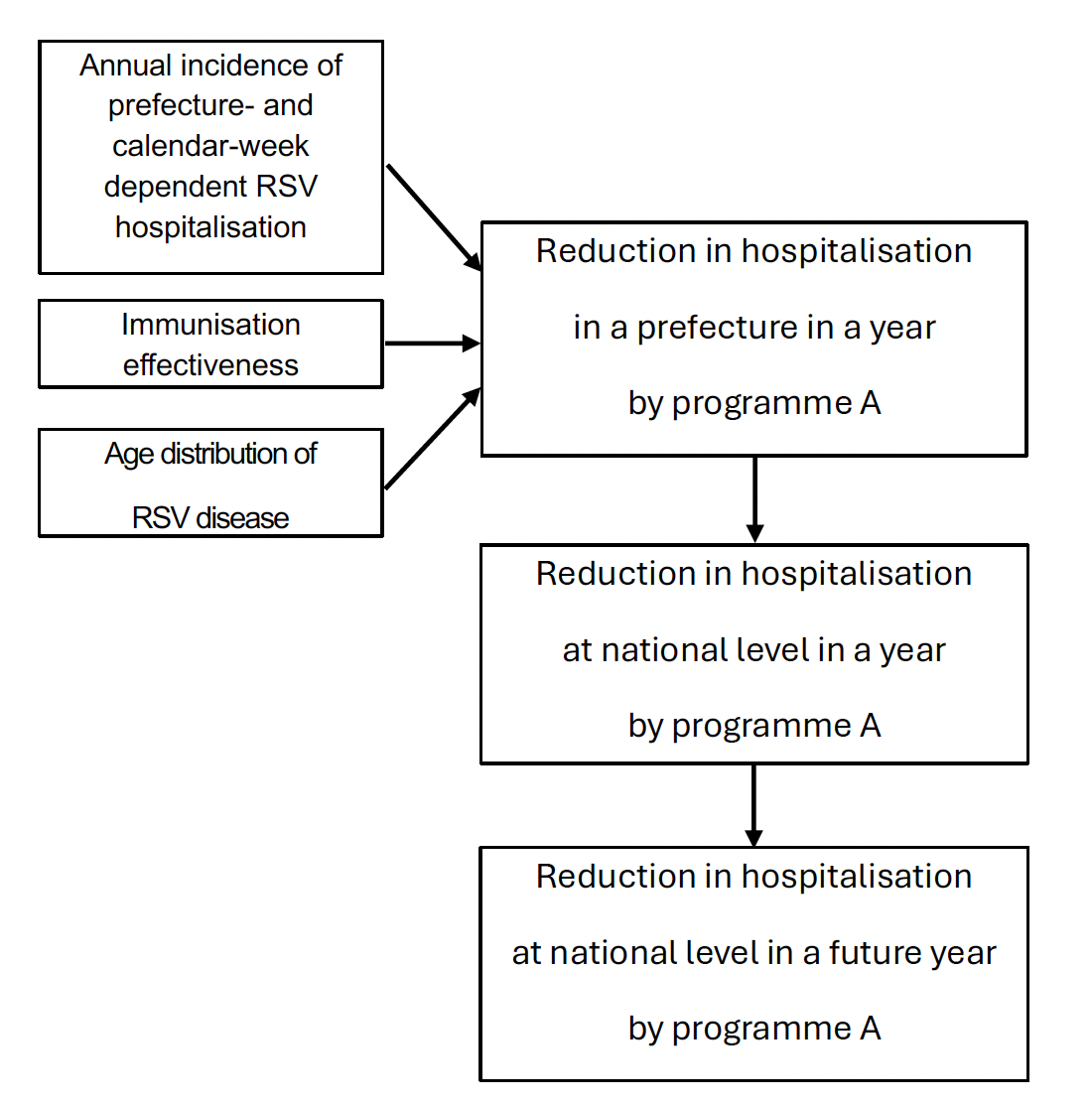

**Figure S1. Overview of the modelling framework**. Our study follows this workflow.

Using this model we calculated reductions in under-1-year-old RSV-associated hospitalisations in a prefecture in a year (*RRpy*) by comparing the numbers of hospitalisaions with and without an intervention as described in Equation (1).

$$RR_{py}$$

$=\sum_{w} \sum_{a} ({Estimated cases}_{w,a} *{Estimated case hospitalisation risk}_{a}* {Estimated effectiveness}_{w,a})$ / $\sum_{w} \sum_{a} ({Estimated cases}_{w,a} *{Estimated hospitalisation risk}_{a})$

= $\sum_{w} \sum_{a} (({{Cases}_{w}*Estimated age dependent proportion}_{a}) *{Estimated case hospitalisation risk}_{a}*{Estimated effectiveness}_{w,a})$ / $\sum_{w} \sum_{a} ({{(Cases}_{w}*Estimated age dependent proportion}_{a})*{Estimated case hospitalisation risk}_{a})$

…Equation (1)

In Equation (1) *w* is calendar week and *a* is age in days. ${Cases}_{w}$refers to the number of RSV cases in calendar week *w* in a study year reported through the Japanese sentinel surveillance [1]. ${Estimated age dependent proportion}_{a}$is estimated proportion of RSV cases at age *a* among under-10-year-old RSV cases, assuming that proportion of RSV incidence from the older age groups is small enough among the pediatric sentinel surveillance data (SI2). We derived RSV case hospitalisation risk among RSV cases at age *a* (${Estimated case hospitalisation risk}_{a}$) from health claims data in 2018 in a study of health claims database (SI2.4) [2]. *Estimated effectiveness* refers to effectiveness of RSVpreF vaccination or nirsevimab administration against RSV hospitalisation for infants aged *a* in week *w*, being estimated using efficacy data from their clinical trials (SI3) [3,4].

To calculate a reduction in RSV hospitalisaions by an intervention across prefectures in a year, we calculated weighted average of reductions as described in Equation (2).

$W_{y}=\sum_{p} ({RR}_{p,y}*{Estimated number of u1y hospitalisation}_{p,y})$ / $\sum_{p} ({Estimated number of u1y hospitalisation}_{p,y}$)

*=* $\sum_{p} ({RR}_{p,y}*\sum_{w} \sum_{a} ({Cases}_{w,p}*{Estimated age dependent proportion}_{a}* {Estimated proportional hospitalisation risk}_{a}))$ /$\sum_{p} \sum_{w} \sum_{a} ({Cases}_{w,p}*{Estimated age dependent proportion}_{a}* {Estimated proportional hospitalisation risk}_{a})$

…Equation (2)

*Wy* is weighted average of reductions in RSV hospitalisations across prefectures in a year *y.* ${RR}_{p,y}$ is multiplied by estimated total number of under-1-year-old RSV hospitalisations in that prefecture in year *y* (${Estimated number of u1y hospitalisation}_{p}$) and divided by the sum of estimated total number of under-1-year-old RSV hospitalisations across prefectures in that year.

#### 2 Estimates of RSV disease burden

##### 2.1 Outline

Since the number of RSV cases in the surveillance data are aggregated in age categories (<6 month old, 6-11 month old, 1, 2, 3, 4, 5, 6, 7, 8, 9 years of age), we estimated the proportion of cases at each age in days by fitting a smoothing function following a gamma distribution to the data.

$A\left( x \right)= \frac{1}{\Gamma(\alpha)\theta^{\alpha}}$ $x^{\alpha-1}℮^{-\frac{\chi}{\vartheta}}$

$A\left( x \right)$ is a function for estimated proportion of cases at age *x* among under-10-year-old RSV cases. We modelled the number of cases in each age category as a cumulative density function of $A\left( x \right)$ in that age category. We jointly fitted the function in the surveillance data by age category and by prefecture.

##### 2.2 Model fitting

The numbers of cases were fitted with a Poisson likelihood using a differential evolution Markov Chain with a snooker updater [5] with 50,000 iterations, then with burn-in of 2000, 10,000 samples were used for the calculation. The fitting was implemented in the R package BayesianTools [6].

| Symbol | Parameter | Prior |
| --- | --- | --- |
| α | Shape of gamma distribution | Uniform(0.1, 10) |
| θ | Scale of gamma distribution | Uniform(0.1, 1000) |

**Table S1. Parameters of age-dependent RSV cases.**

##### 2.3 Fitted results

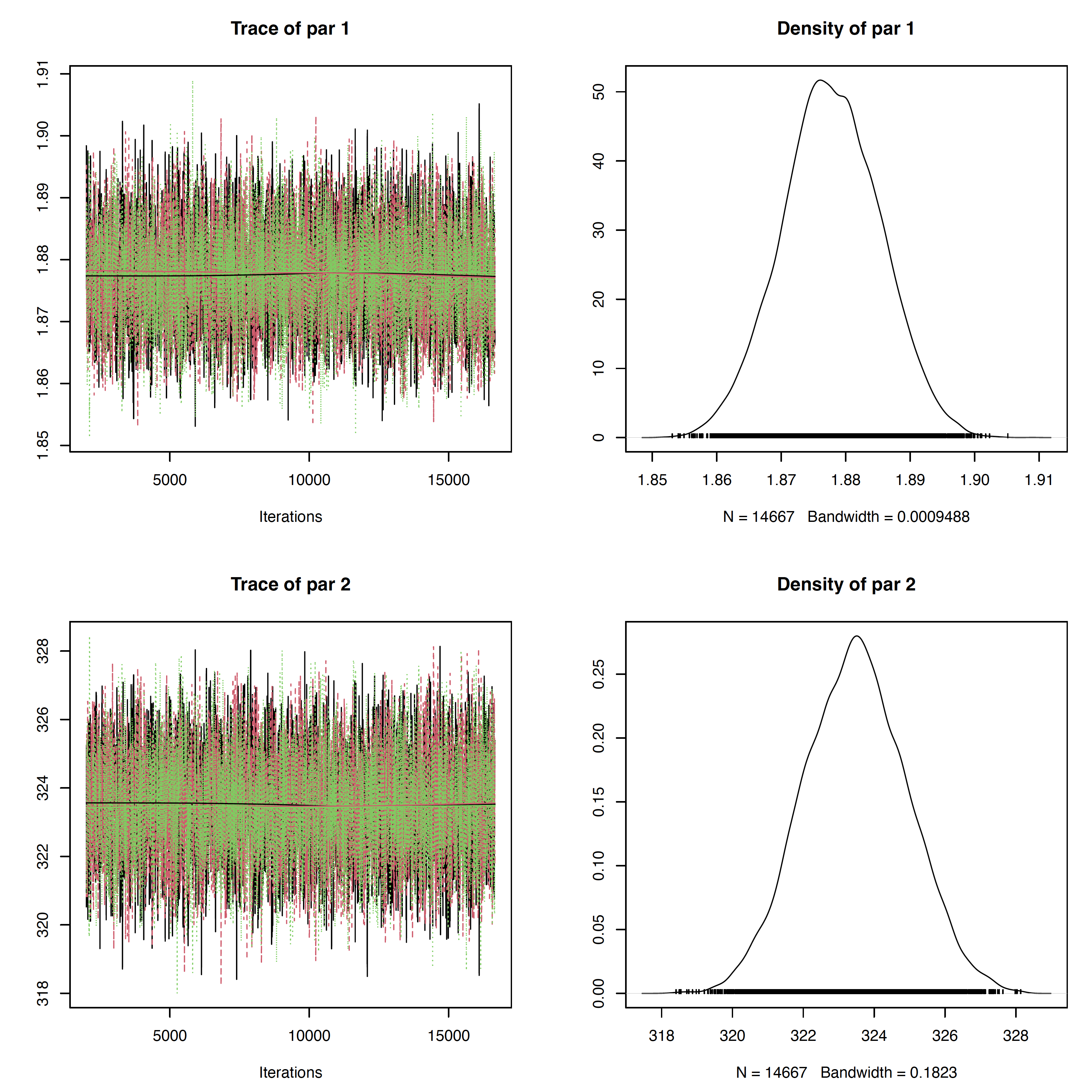

**Figure S2. Trace plots (left) and density plot (right) of fitted model by parameter.** Each parameter corresponds to the parameters in Table S1.

| Symbol | Parameter | ESS |
| --- | --- | --- |
| α | Shape of gamma distribution | 5170 |
| θ | Scale of gamma distribution | 5203 |

**Table S2. Effective sample size (ESS) for** **parameters of age distribution of RSV disease.**

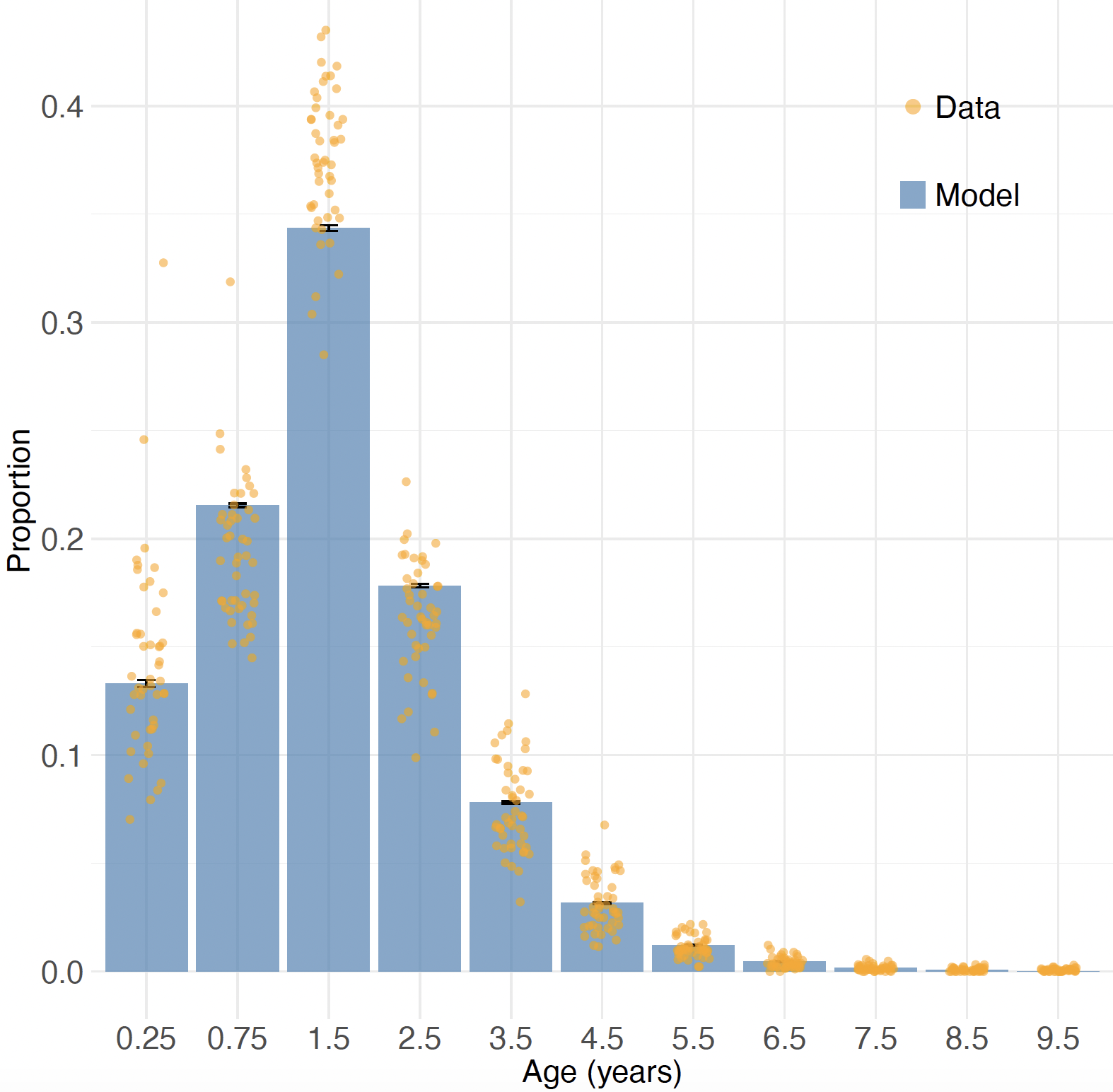

**Figure S3. Proportion of RSV cases in each age category among under-10-year-old RSV cases.** Orange dots show the proportion of reported RSV cases in that age category (0 months to 5 months, 6 months to 11 months, 1 year, 2 years, 3 years, 4 years, 5 years, 6 years, 7 years, 8 years, 9 years) by prefecture through the sentinel surveillance system. Blue bars show the estimated proportion by age category.

##### 2.4 Case hospitalisation risk

To estimate age-dependent RSV-associated case hospitalisation risk among RSV medically-attended cases, we referred to a study based on a health claim database [2]. The Japanese Medical Data Center database provides anonymized inpatient, outpatient, and pharmacy claims from health insurance associations in Japan [2]. We derived the numbers of RSV patients for “All RSV patients” and “Inpatients” by age in months in 2018 by digitizing Fig. 1. Then we fitted a generalized additive model with a smoothing spline to the case hospitalisation risk.

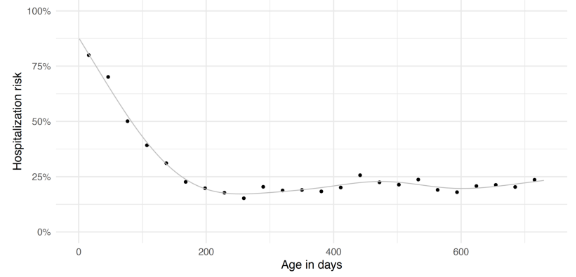

**Figure S4. RSV case hospitalisation risk among RSV patients during the first two years of life.** Black dots show the observed RSV case hospitalisation risk from [2] and gray line shows the modelled risk.

#### 3 Estimates of waning efficacy

##### 3.1 Outline

Observed disease outcomes in the trials suggested that vaccine efficacy wanes after birth [3,4]. We modelled VE_est(t) vaccine efficacy at time *t* after birth as the product of *V0*, the initial value of efficacy, and *w(t)*, the function of protection decay of efficacy after birth. We assumed that *w(t)* follows an Erlang-2 distribution.

$w\left( t \right)= \sum_{n=0}^{k-1} \frac{{T\_v}^{n}t^{n}e^{-T\_v * t}}{n!}$... Equation (3)

Equation (3) represents the counter cumulative density function of Erlang-2 distribution.

We estimated the efficacy waning rate by fitting the modelled outcomes to the observed outcomes in the trial. The incidence in the intervention arm at time t after birth I_v(t) was estimated using modelled efficacy and the incidence in the placebo arm I_p(t).

*I_v (t) = I_p(t)**(1 - *VEest(t)*)

Since the trial outcomes were grouped into 30-day intervals from birth to 180 days after birth, the means of the estimated vaccine efficacy during 30-day period were used to calculate the number of infected individuals in intervention arm *I_v (t)* during that period. The modelled outcomes *C* (i.e., individuals newly counted as that disease outcome during that period) in the intervention arm were fitted to the RSV disease outcomes in the trial.

*C* *~ B*(*N*, *I_v (t_mid)*)

In the intervention arm*, N* is the total number of individuals in the arm, and *I_v (t_mid)* is the incidence in the intervention arm in the middle of that period assuming incidence is constant during that period. We assume that efficacy against RSV disease outcomes with different severities wanes at the same rate but the initial values *V0* are severity-dependent.

##### 3.2 Overview of data

We used the outcome of the phase 3 trial of RSVpreF [4] and the phase 2b and 3 trials of nirsevimab [3] (Table S3). We calculated the incremental efficacy of RSVpreF from data on cumulative counts of severe RSV-Positive MA-LRTIs (Table S3A) and less severe RSV-Positive MA-LRTIs (Table S3B) from 0 to 180 days after birth by trial arm.

For nirsevimab, we calculated incremental efficacy of long-acting monoclonal antibody from a pooled efficacy analysis of clinical trials: the phase 2b trial enrolled otherwise healthy infants born preterm (gestational age from 29 to less than 35 weeks) and phase 3 trial enrolled healthy infants born at term or late preterm (gestational age 35 weeks or older).

The data used are the number at risk and the number censored at each time point for first medically attended RSV LRTI (Table S4A) and first hospital admission for medically attended RSV LRTI (Table S4B) from 0 to 150 days post-dose by trial arm.

(A) Cumulative counts of severe RSV-positive MA-LRTIs through 180 days of age

| Post-dose time (days) | RSVpreF 120μg  N = 3585 | Placebo  N = 3563 |
| --- | --- | --- |
| 0 - 30 | 1 | 10 |
| 30 – 60 | 4 | 28 |
| 60 – 90 | 6 | 34 |
| 90 - 120 | 13 | 49 |
| 120 - 150 | 18 | 61 |

(B) Cumulative counts of less severe RSV-positive MA-LRTIs through 180 days of age

| Post-dose time (days) | RSVpreF 120μg  N = 3585 | Placebo  N = 3563 |
| --- | --- | --- |
| 0 - 30 | 2 | 15 |
| 30 – 60 | 14 | 38 |
| 60 – 90 | 25 | 59 |
| 90 - 120 | 40 | 88 |
| 120 - 150 | 55 | 110 |

**Table S3. Cumulative number of maternal vaccine trial outcomes by 30-day interval from 0 to 180 days after birth regarding (A) severe RSV-Positive MA-LRTIs and (B) less severe RSV-Positive MA-LRTIs from the phase 3 trial of maternal vaccine [4].**

(A) Outcomes for first medically attended RSV LRTI

| Post-dose days | Nirsevimab  Number at risk (number censored) | Placebo  Number at risk (number censored) |
| --- | --- | --- |
| 0 | 1564 (0) | 786 (0) |
| 30 | 1553 (8) | 772 (6) |
| 60 | 1546 (11) | 756 (7) |
| 90 | 1538 (17) | 737 (8) |
| 120 | 1527(21) | 729 (9) |
| 150 | 1519 (1545) | 724 (735) |

(B) Outcomes for first hospital admission for medically attended RSV LRTI

| Post-dose days | Nirsevimab  Number at risk (number censored) | Placebo  Number at risk (number censored) |
| --- | --- | --- |
| 0 | 1564 (0) | 786 (0) |
| 30 | 1554 (8) | 778 (6) |
| 60 | 1547 (11) | 769 (7) |
| 90 | 1540 (17) | 761 (8) |
| 120 | 1535 (21) | 757 (10) |
| 150 | 1529 (1555) | 753 (765) |

**Table S4. Nirsevimab trial outcomes by 30-day interval from 0 to 150 days after birth regarding (A) first medically attended RSV LRTI and (B) first hospital admission for medically attended RSV LRTI [3].**

##### 3.3 Model fitting

For either RSVpreF or nirsevimab, the numbers of counts of the two outcomes with different severity were fitted with a binomial likelihood. For RSVpreF, the Erlang-2 model was fitted using a Metropolis-Hastings sampler in MCMC with 100,000 iterations and thinned by 10, then 10,000 samples were used for the calculation.

For nirsevimab, the model was fitted using a differential evolution Markov Chain with a snooker updater [5] with 100,000 iterations with burn-in of 2,000 and thinned by 5, then 10,000 samples were used for the calculation. Both fittings were implemented in the R package BayesianTools [6].

| Symbol | Parameter | Prior |
| --- | --- | --- |
| VE0_s | Vaccine efficacy against severe disease at birth | Uniform(0.001, 0.999) |
| VE0_l | Vaccine efficacy against less severe disease at birth | Uniform(0.001, 0.999) |
| T_v | Rate of Erlang-2 distribution | Uniform(0.001, 0.999) |

**Table S5. Parameters of waning efficacy of RSVpreF.**

| Symbol | Parameter | Prior |
| --- | --- | --- |
| VE0_s | Efficacy against first hospital admission for medically attended RSV LRTI | Uniform(0.001, 0.999) |
| VE0_l | Efficacy against first medically attended RSV LRTI | Uniform(0.001, 0.999) |
| T_v | Rate of Erlang-2 distribution | Uniform(0.001, 0.999) |

**Table S6. Parameters of waning efficacy of nirsevimab.**

##### 3.4 Fitted results

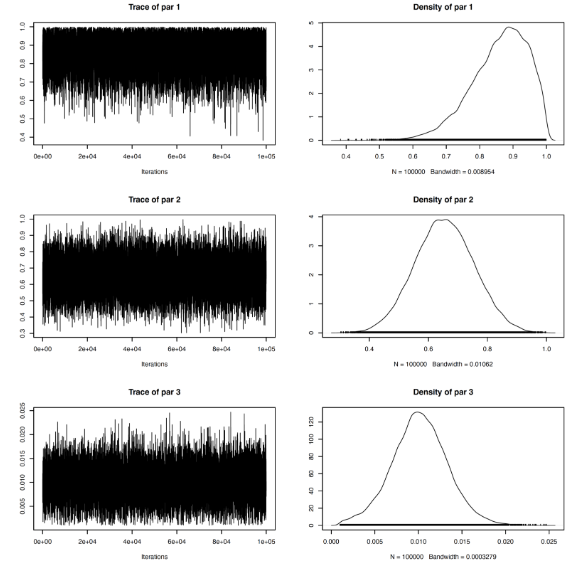

**Figure S5 Trace plots (left) and density plot (right) of fitted model by parameter.** Each parameter corresponds to the parameters in Table S5.

| Symbol | Parameter | ESS |
| --- | --- | --- |
| VE0_s | Vaccine efficacy against severe disease at birth | 6565 |
| VE0_l | Vaccine efficacy against less severe disease at birth | 7231 |
| T_v | Rate of Erlang-2 distribution | 6585 |

**Table S7. Effective sample size (ESS) for** **parameters of waning efficacy of RSVpreF.**

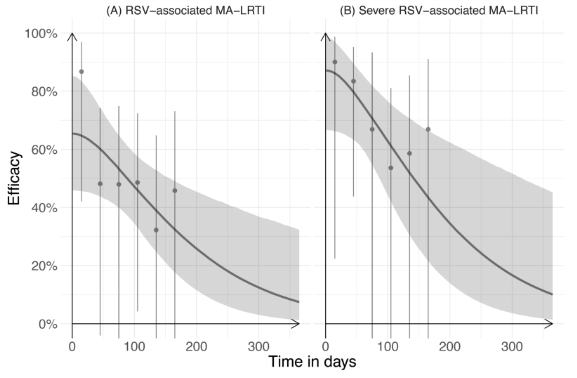

**Figure S6.** **Efficacy of RSVpreF against RSV-associated (A) less severe and (B) severe MA-LRTIs during the first year of life.** Efficacy against RSV-associated severe MA-LRTIs observed in the trial is shown as gray dots together with binomial 95% confidence intervals. Modelled efficacy is shown as a gray line with gray-shaded 95% credible intervals.

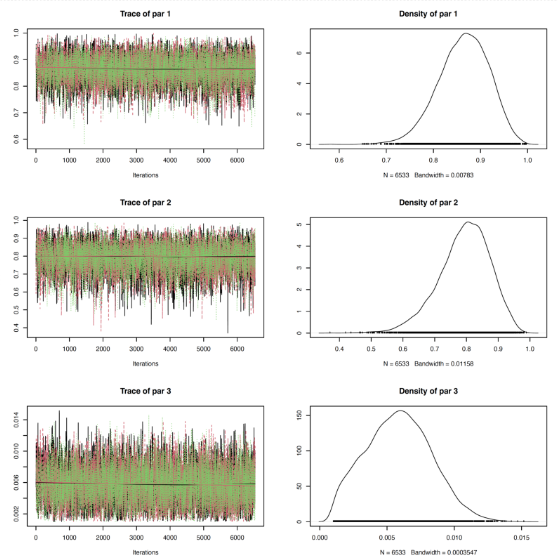

**Fig S7. Trace plots (left) and density plot (right) of fitted model by parameter.** Each parameter corresponds to the parameters in Table S6.

| Symbol | Parameter | ESS |
| --- | --- | --- |
| VE0_s | Efficacy against first hospital admission for medically attended RSV LRTI | 7266 |
| VE0_l | Efficacy against first medically attended RSV LRTI | 7238 |
| T_v | Rate of Erlang-2 distribution | 7578 |

**Table S8. Effective sample size (ESS) for** **parameters of waning efficacy of nirsevimab.**

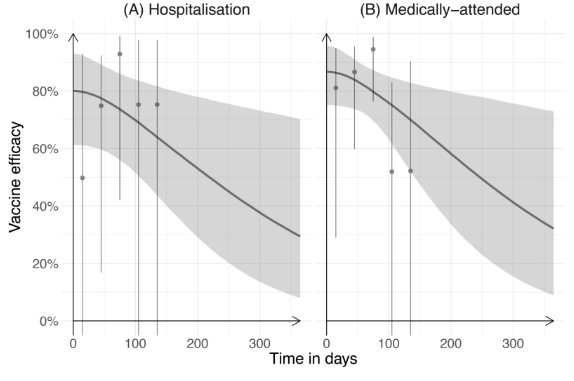

**Fig S8.** **Efficacy of nirsevimab against RSV-associated (A) hospitalisation and (B) medically-attended infection during the first year of life.** Efficacy observed in the trial is shown as gray dots together with binomial 95% confidence intervals. Modelled efficacy is shown as a gray line with gray-shaded 95% credible intervals.

#### 4 Intervention model

##### 4.1 Outline

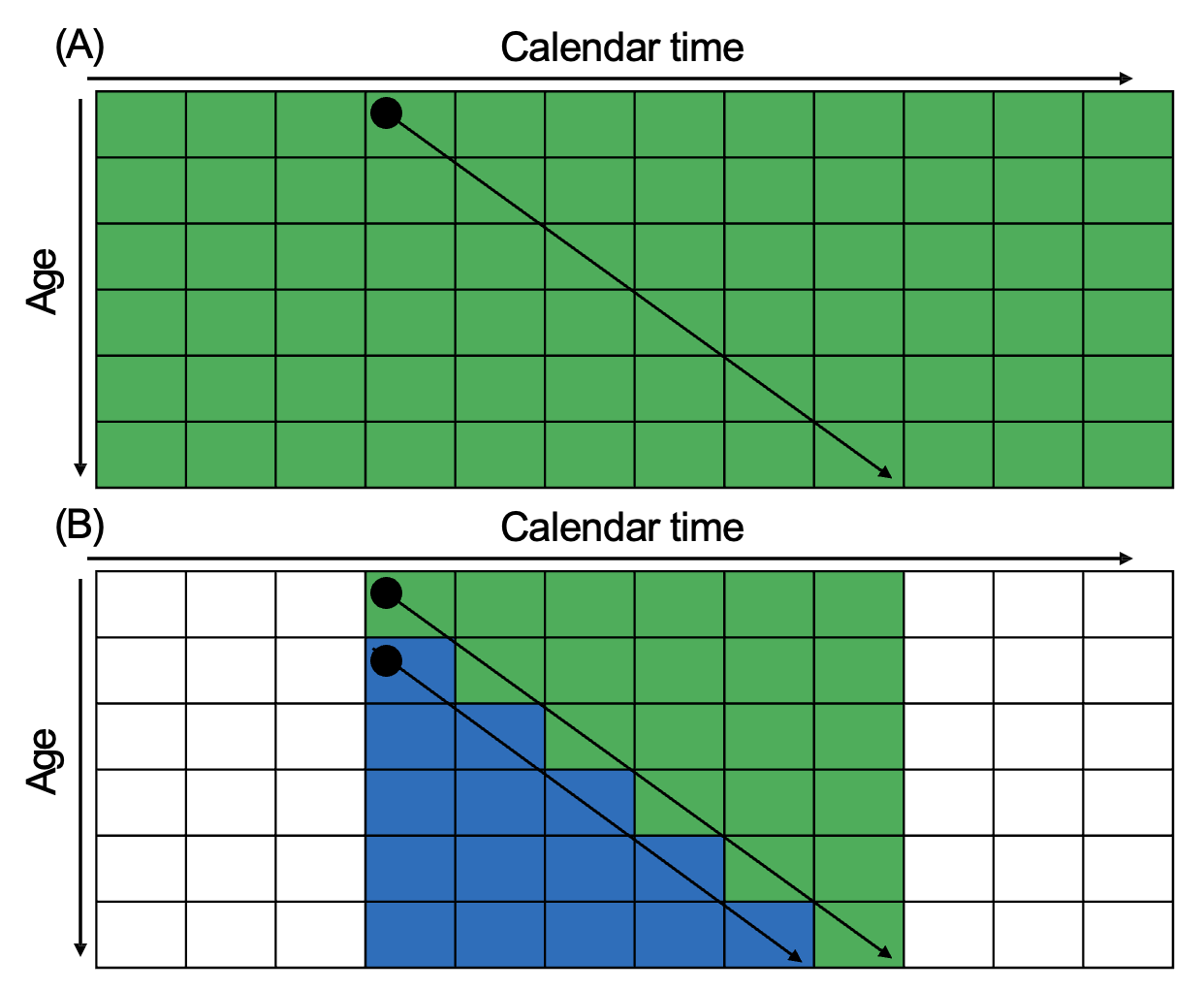

**Fig S9 Schema of RSV infant immunization programs: (A) year-round administration and (B) seasonal administration combined with catch-up administration of nirsevimab.** Horizontal arrows show calendar time and vertical arrows show age of infants. Black dots show time points when infants are born to mothers who have received RSVpreF or when they receive nirsevimab. Diagonal arrows show aging of infants. The colored area in green shows that infants are immunised as birth cohort (i.e., infants are born to mothers who have received RSVpreF or receive nirsevimab at birth) and the colored area in blue shows that infants are immunised as catch-up cohort at the beginning of their first RSV season.

##### 4.2 Immunisation status

We defined the content of immunisation an infant who is in calendar week *i* has received based on age in days *a* of that infant.

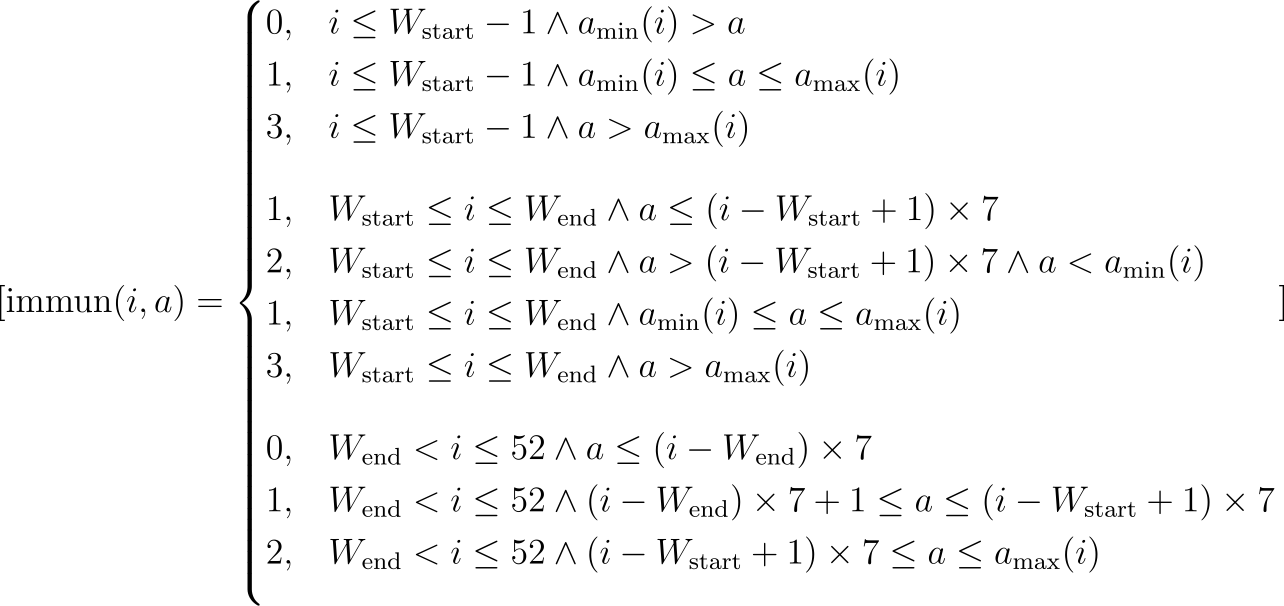

Immunisation status is categorised as non-immunised (0), immunised as birth cohort (1), immunised this year as catch-up cohort (if there is a catch-up administration) (2), or immunised in the previous year as catch-up cohort (if there was a catch-up administration) (3). $W_{start}$ and $W_{end}$ is the start and end week of immunisation.

$a_{min}(i)$ and $a_{max}(i)$ are defined to model protection from immunisation from the previous year. In this study we assumed that infants who received immunisation in the previous year will not receive immunisation in the current year.

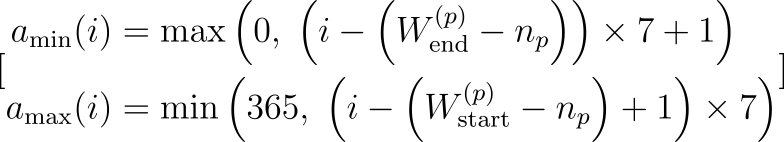

$n_{p}$ is the number of weeks in the previous year.

##### 5. Influence on duration of immunity on additional benefit of seasonal and catch-up programme

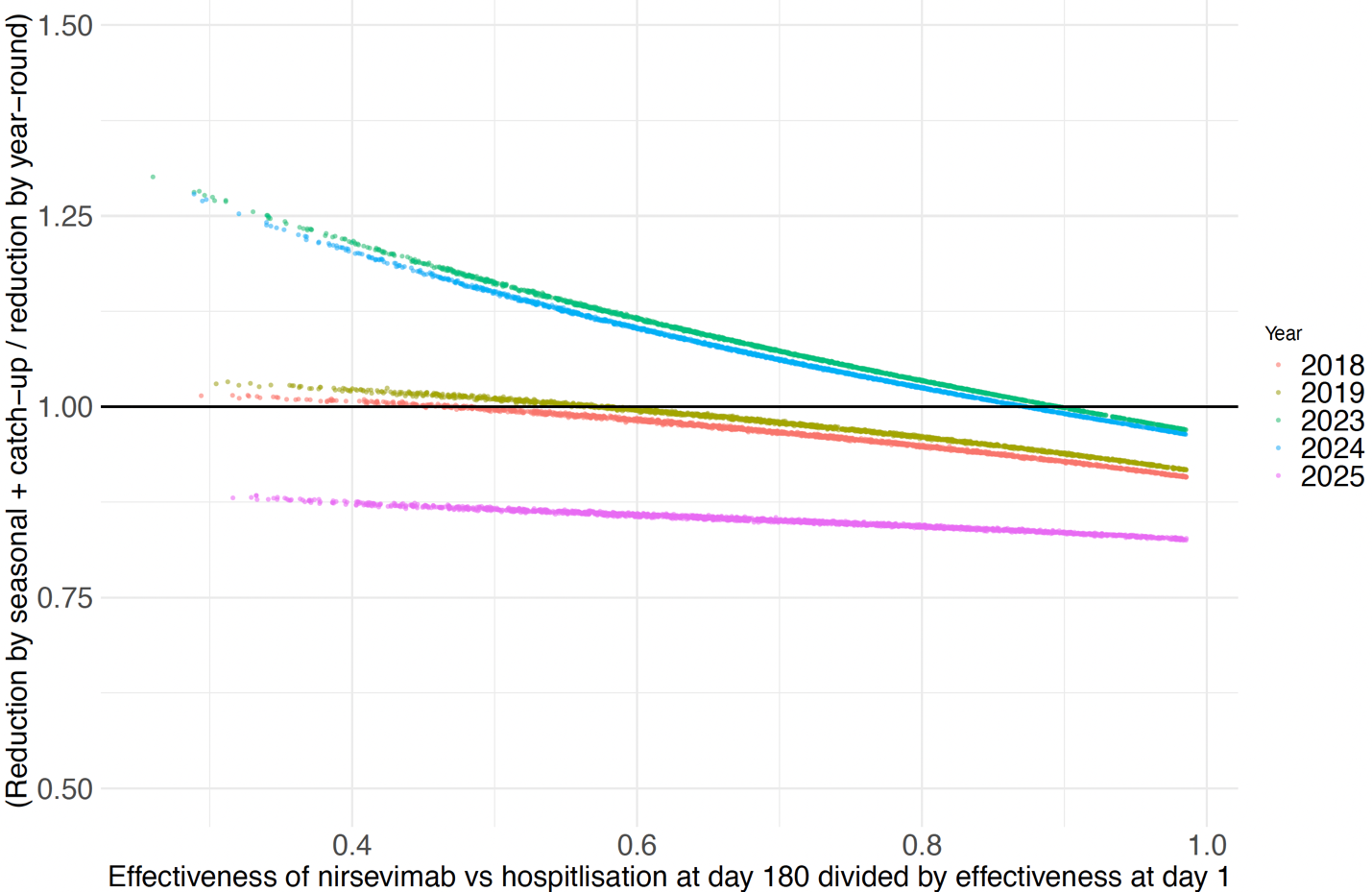

**Figure S10 Relationship between estimated waning effectiveness of nirsevimab and reductions in RSV hospitalisaions by nirsevimab-based prefecture-specific seasonal and catch-up programme in baseline scenario compared to the year-round programme.** The x axis shows the ratio between the effectiveness of nirsevimab in 180 days after administration compared to the effectiveness at birth. The y axis shows ratio between reductions from seasonal and catch-up programme compared to reduction from year-round programme.

##### 6. Nationally synchronised seasonal administration

(A)

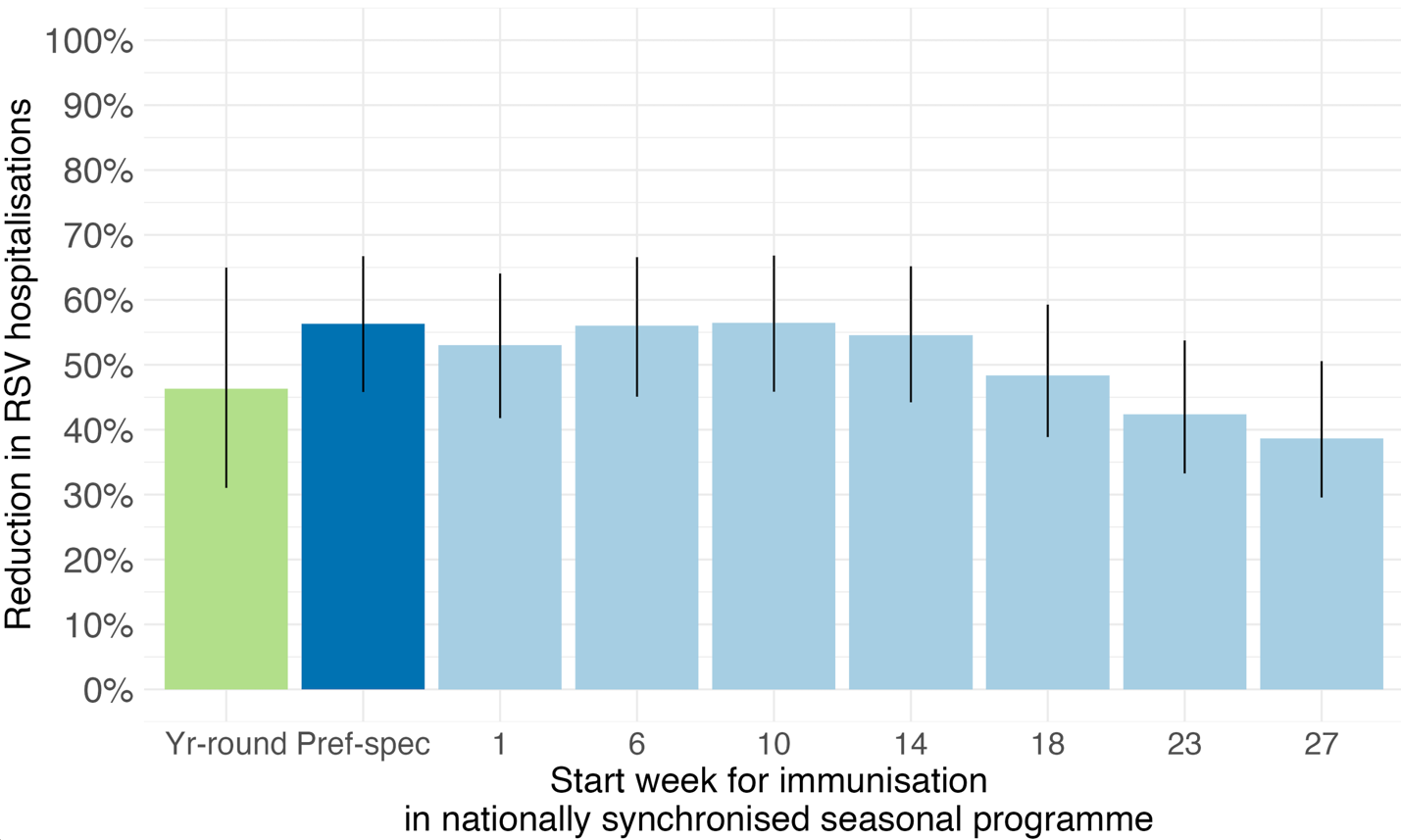

(B)

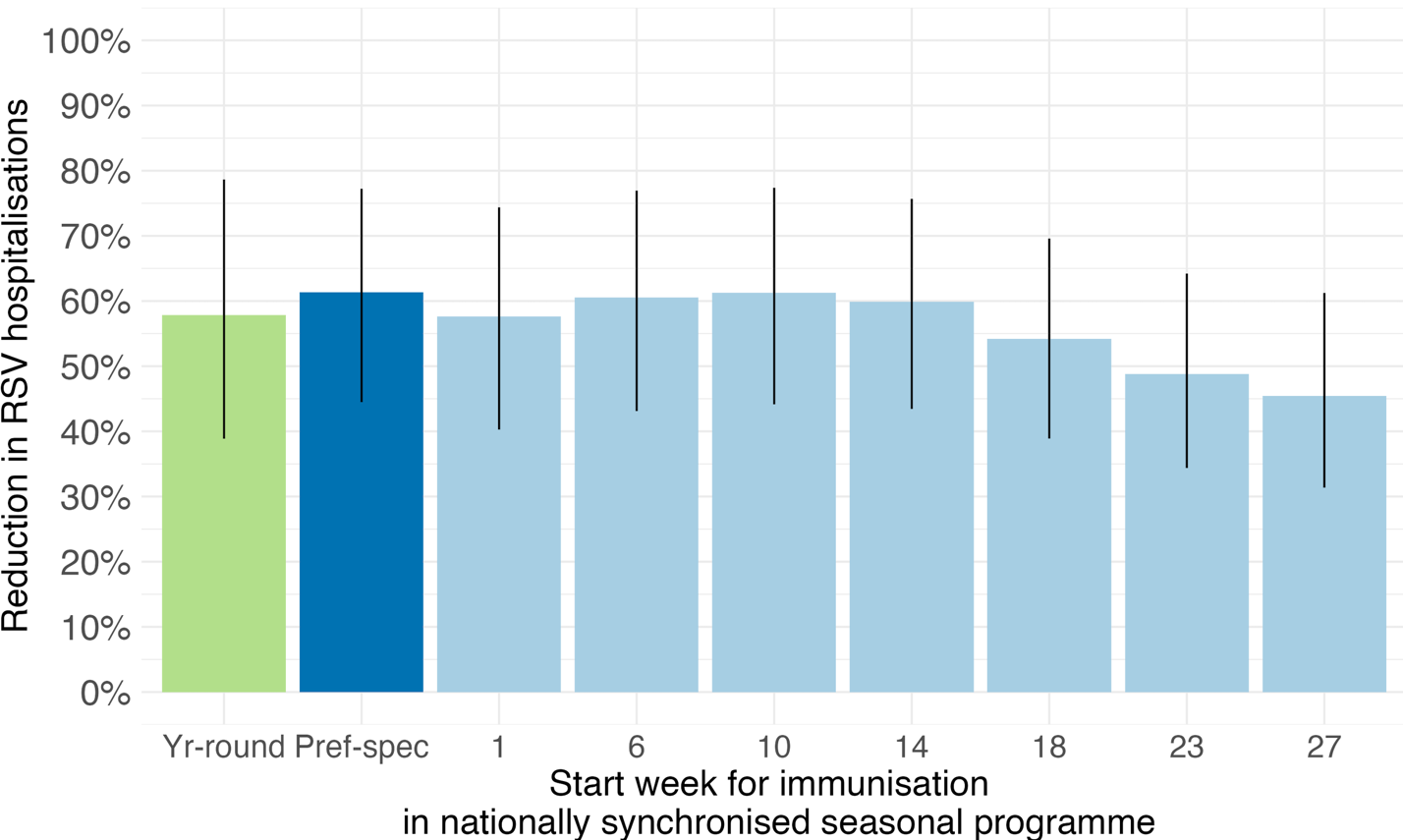

**Figure S11 Comparison of estimated reductions in RSV infant immunisation between programmes from the status quo.** Estimated reductions in RSV hospitalisations by programmes for which RSVpreF is used for infants born during RSV season (A) and by programmes for which nirsevimab is used for infants born during RSV season (B). Green bars show redctions by year-round programmes, thick blue bars show reductions by seasonal and catch-up programmes with prefecture-specific seasonal immunisation timing, and light blue bars show reductions by seasonal and catch-up programmes with the same timing for all prefectures (nationally synchronised seasonal programme). Reductions by nationally synchronised seasonal programme are estimated by varying start timings in all prefectures. The starting weeks (1, 6, 10, 14, 18, 23, 27) correspond to calendar months from January to July.

##### 7. Regional heterogeneity in impact under nationally synchronized seasonal administration

Assuming the future RSV incidence would be the same as one of the four study years (2018, 2019, 2023, 2024, and 2025), we find that the overall impact under nationally synchronised seasonal and catch-up programmes staring in week 10 (as one of the best timings) is a reduction in RSV hospitalisation of 52% (95%UR: 38%, 65%) for RSVpreF and 57% (95%UR: 39%, 75%) for nirsevimab. The prefecture level impact ranged from a reduction of 45% (95%UR: 32%, 60%) and 51% (95%UR: 33%, 70%) in Hokkaido to a reduction of 59% (95%UR: 48%, 69%) and 63% (95%UR: 45%, 79%) in Okinawa, respectively. We estimated that proportionally the RSVpreF-based prefecture-specific seasonal administration strategy would be better in reducing RSV infant hospitalisations by 1.04 times in Tottori, while it would decrease the reduction by 0.97 times in Okinawa (Fig S12A). The nirsevimab-based strategy would increase the reduction by 1.04 times in Yamagata and decrease by 0.98 times in Kagoshima (Fig S12B).

(B)

(A)

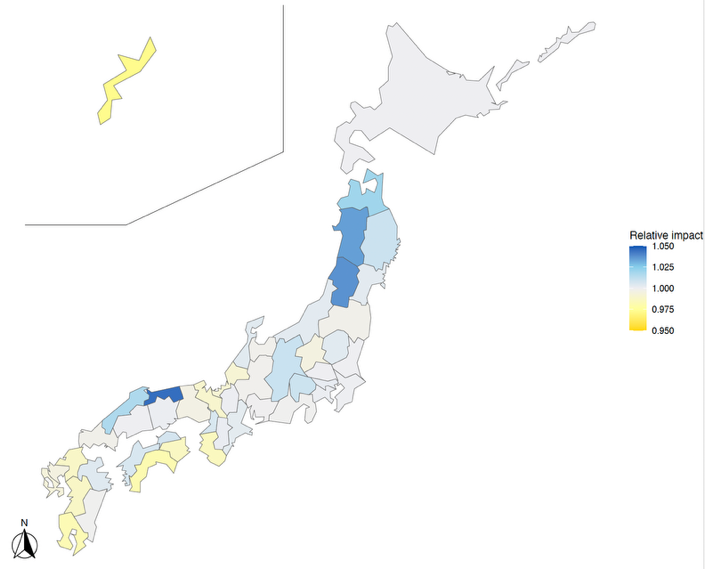

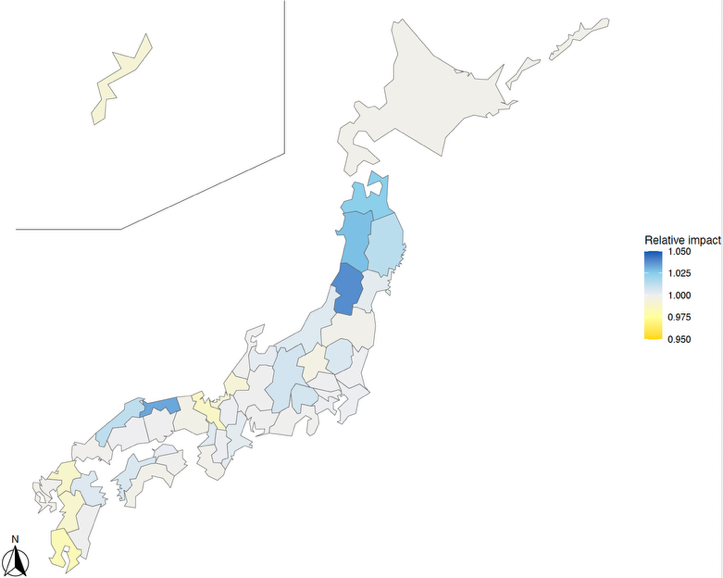

**Figure S12 Additional benefits of prefecture-specific strategy of seasonal administration combined with catch-up administration of nirsevimab for infants born outside the RSV season, compared to nationally synchronised strategy starting in week 10 in the baseline scenario.** The estimates are based on RSV surveillance data in 2018, 2019, 2023, 2024 and 2025. (A) RSVpreF is used for infants born during RSV season (B) nirsevimab is used for infants born during RSV season.
